# Investigation of the ventilation situation in a lecture room of building 033 at the Universität der Bundeswehr München

**DOI:** 10.1101/2021.03.17.21253800

**Authors:** Christian J. Kähler, Thomas Fuchs, Rainer Hain

## Abstract

The SARS-CoV-2 pandemic is limiting both the private and public lives of many people around the world. It is now considered certain that SARS-CoV-2 is transmitted via droplets, smear infection, and aerosol particles. While simple masks, spacing, and hand hygiene significantly reduce the risk of infection via the first two routes mentioned, the risk from aerosol particles remains. These small particles move with the air in the room and spread unhindered throughout it. To reduce the risk of infection from viruses present in aerosol particles, the following options exist. First, good respiratory masks can be worn to reduce the viral load in the inhaled air. Another option is to make the viruses harmless (e.g., by UV light). A third option is to reduce the viral load in the room by bringing in virus-free air and moving contaminated air out or cleaning the air in the room. To investigate how well virus load reduction via ventilation works in a real lecture room, measurements were carried out at the Universität der Bundeswehr München (University of the Federal Armed Forces Munich). The lecture room holds a maximum of approx. 90 people and has a ventilation system as well as 2 windows that can be opened. In the absence of a ventilation system in a comparable room, the effectiveness of a room air cleaner was also investigated.

## 1. Introduction

The pandemic caused by SARS-CoV-2 is currently spreading worldwide. According to the current state of research, it is well accepted that the transmission of the infection occurs by droplet infection, by aerosol particles contaminated with viruses and with minor likelihood also by smear infection. The risk of droplet infection can be reduced by spacing people apart and wearing masks (Pringle et al. (2020)). The risk of smear infection can be reduced primarily by regular thorough hand hygiene and avoiding touching mucous membranes (Centers for Disease Control and Prevention (2021a)). In contrast, aerosol particles contaminated with viruses spread undisturbed and can also be inhaled over greater distances (Editorial The Lancet Respiratory Medicine (2020)). A reduction of the inhaled and possibly virus-laden aerosol particles can be achieved by a) a suitable respiratory protection mask, b) a reduction of the virus concentration, or c) reducing the residence time in a contaminated room.

A respirator mentioned in point a) must be capable of filtering aerosol particles with a sufficiently high efficiency. This requires an N95/KN95/FFP2 standard or better (Gesellschaft für Aerosolforschung (2020)). Simple mouth-nose covers and surgical masks do not have sufficient filtration efficiency (Kähler and Hain (2020)). In addition to the lack of filtering efficiency of these coverings and masks, a major problem is the gap between the mask and the face through which the aerosol particles are inhaled unfiltered (Kähler and Hain (2020)).

The reduction of the virus concentration described under point b) can be achieved by various mechanisms. For example, via inactivation of the viruses by means of UV radiation, electrical charges, chemical processes or separation of the viruses/particles by filters. Likewise, a reduction is achieved by bringing uncontaminated air into a room and taking out the contaminated room air. This can be achieved by ventilation through windows or - if available - by a heating, ventilation and air conditioning (HVAC) system. Finally, the stationary virus concentration can be effectively reduced with mobile room air cleaners with suitable filters (Bluyssen et al. (2020); Curtius et al. (2020); Kähler at al. (2020a); Kähler et al. (2020b); Küpper et al. (2020)).

In the context of conducting classroom teaching at times of the coronal pandemic, the question arises as to how well the various methods for reducing the viral load work in practice. This is important because an efficient and lasting reduction of possible viruses in rooms lowers the risk of infection or enables a longer duration of stay until an infection takes place, see Müller et al. (2020). To answer the question, experiments were conducted in a real auditorium at the Universität der Bundeswehr München, Germany. The lecture rooms at the ends of building 033 have modern HVAC systems and two windows on opposite sides, to generate mixed and displacement ventilation. The number and size of the windows fulfills the state regulations. For comparison, the efficiency of shock and cross ventilation was also analyzed under the temperature and wind conditions that prevailed on the measurement day. Furthermore, the efficiency of a mobile room air cleaner was investigated as well, since these devices are used more and more in the pandemic and since they are recommended by many groups and organizations, e.g. Harvard T.H. Chan School of Public Health (2021), Centers for Disease Control and Prevention (2021b); Deutsche Physikalische Gesellschaft e.V. (2021).

## 2. Measurement setup and data analysis

The measurements were carried out in lecture room 2431 in building 033 of the Universität der Bundeswehr München (University of the Federal Armed Forces in Munich). A panoramic view of the lecture room is shown in Fig. 1. The supply and exhaust air of the HVAC flows into and out of the room via a total of 12 openings in the ceiling. A sketch of the auditorium floor plan and the measurement location are shown in Fig. 2. The two windows each have an opening of 695^W^ mm x 1145^H^ mm.

**Figure 1:**
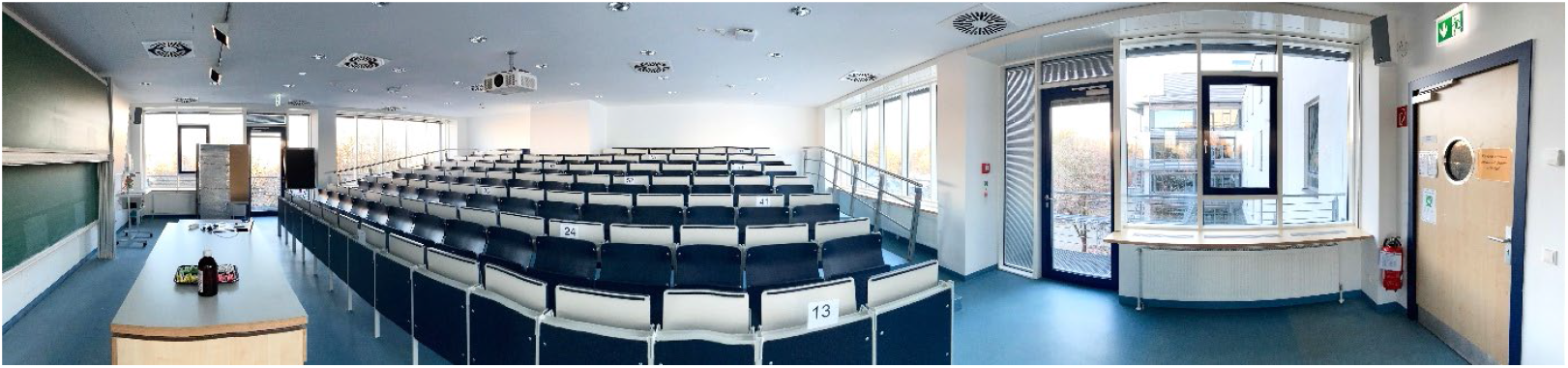
Panoramic view (distorted) of lecture room 2431 in building 033.

**Figure 2:**
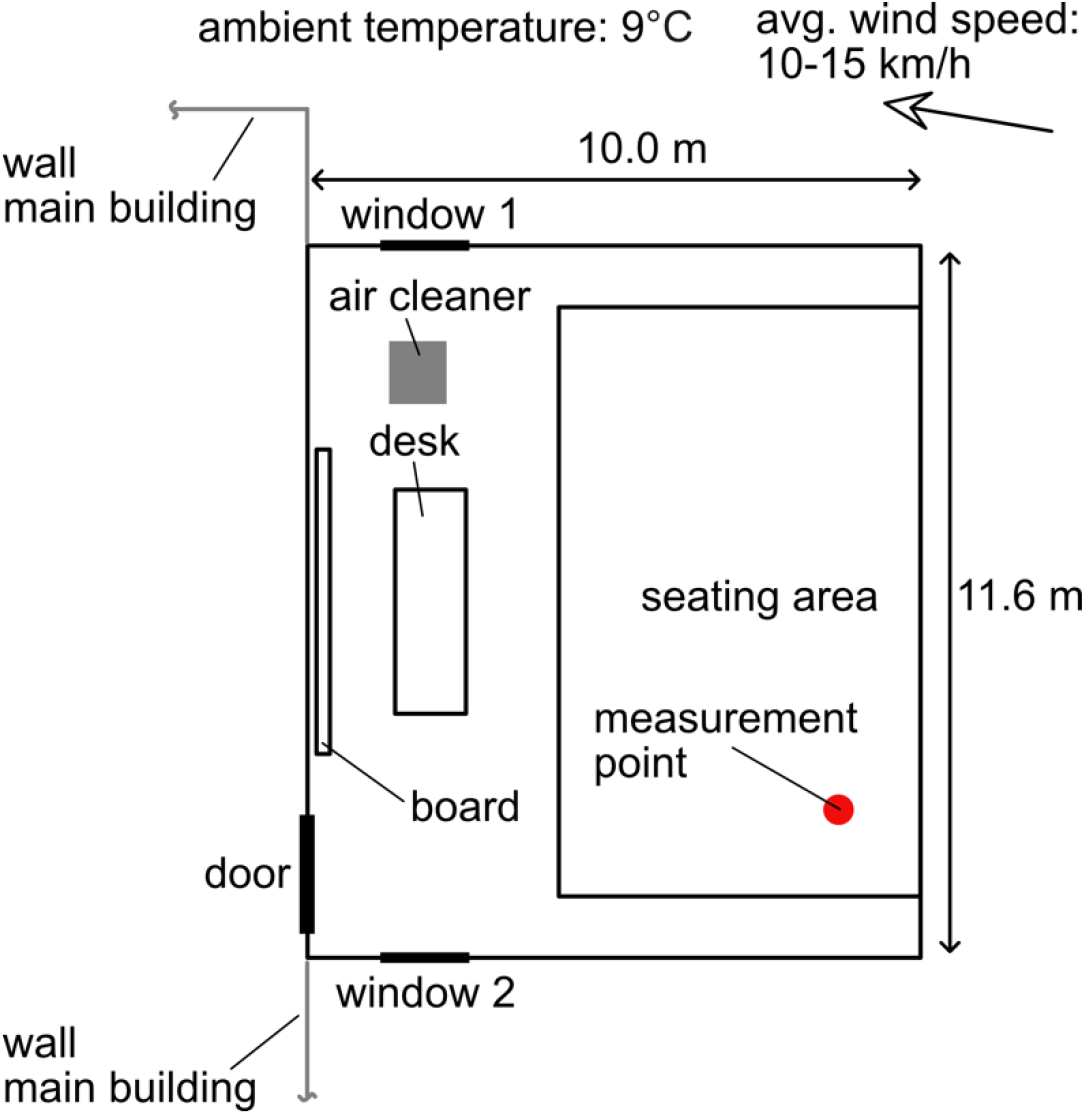
Sketch of the lecture room 2431 in building 033 with the overview of lecture room dimensions and the position of the air cleaner and the measurement location. The size of each window opening with fully opened window is 695^W^ mm × 1145^H^ mm.

To determine the cleaning efficiency, artificially generated aerosol particles of DEHS (average diameter approx. 0.4 µm) were introduced into the room and homogeneously distributed before the start of the measurement. The size of these particles matches the range of particle sizes emitted by infected humans and contaminated with viruses. The particles follow the flow in the room almost ideally and the settling due to gravity is negligible.

The particle concentration is recorded over time using the Promo 3000 particle counter with the Welas 2300 sensor head (Palas GmbH, Germany).

The decay of the particle concentration is measured and fitted with an exponential function. From the function, the decay rate *k* with the unit [1/h] is determined to estimate the efficiency of the room air cleaning. In ventilation technology, *k* is also known as air exchange rate. With the aid of *k*, the development of the particle concentration *c* over time can be determined, provided that the room volume *V* [m^3^] and the strength *S* [particles/h] of the source of contamination are known. The concentration *c*_steady-state_, which becomes stationary after a longer period of time, can be calculated as follows:

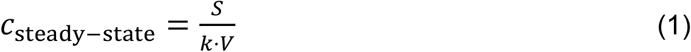

The higher *k* is, the faster potentially dangerous aerosol particles will be removed, or the lower the concentration will be after a longer period of time.

## 3. Results

The measured, normalized particle concentrations over time are displayed in Figs. 3 and 4. The different decreases of the aerosol particle concentrations indicate that the room air cleaning performance over time differs quite strongly.

**Figure 3:**
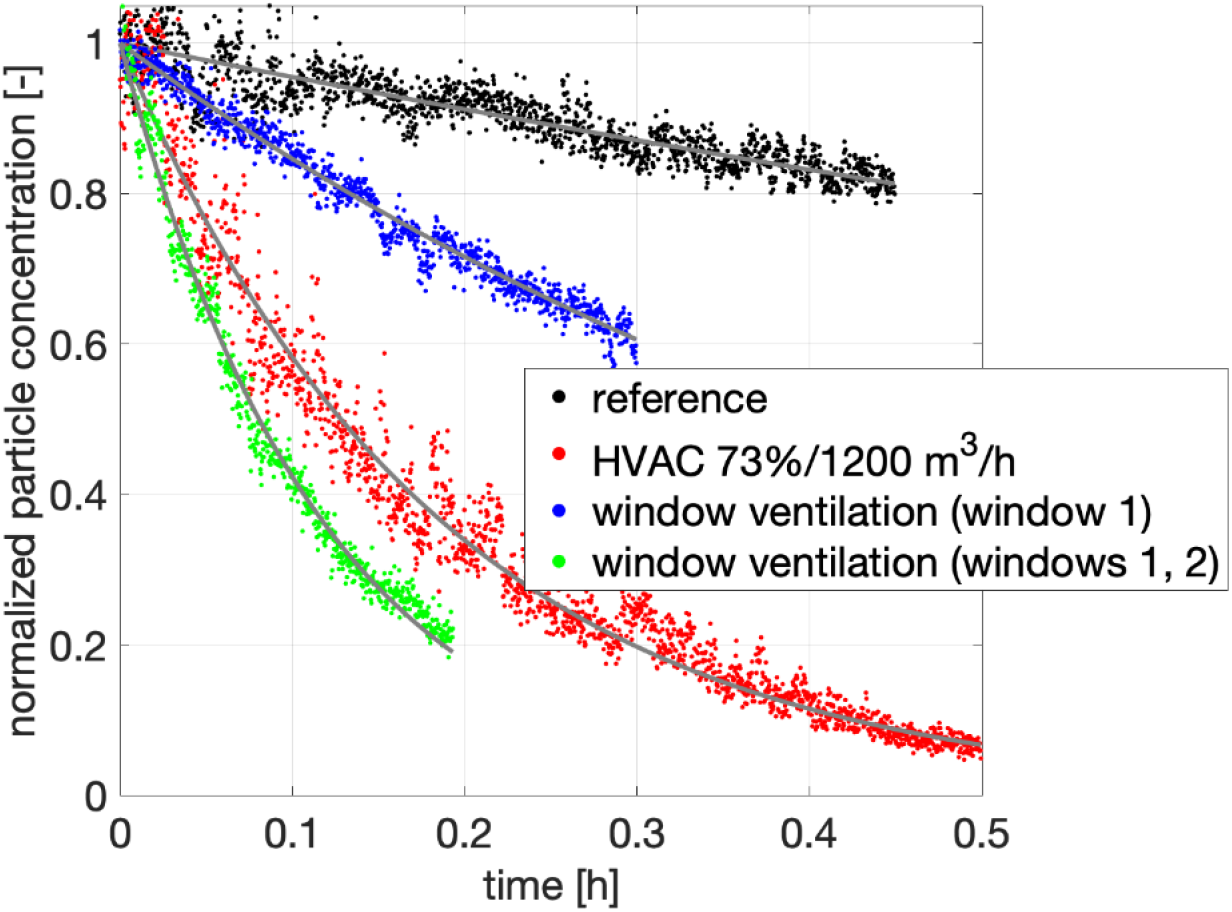
Normalized particle concentrations over time for the reference measurement, the HVAC system and window ventilations.

**Figure 4:**
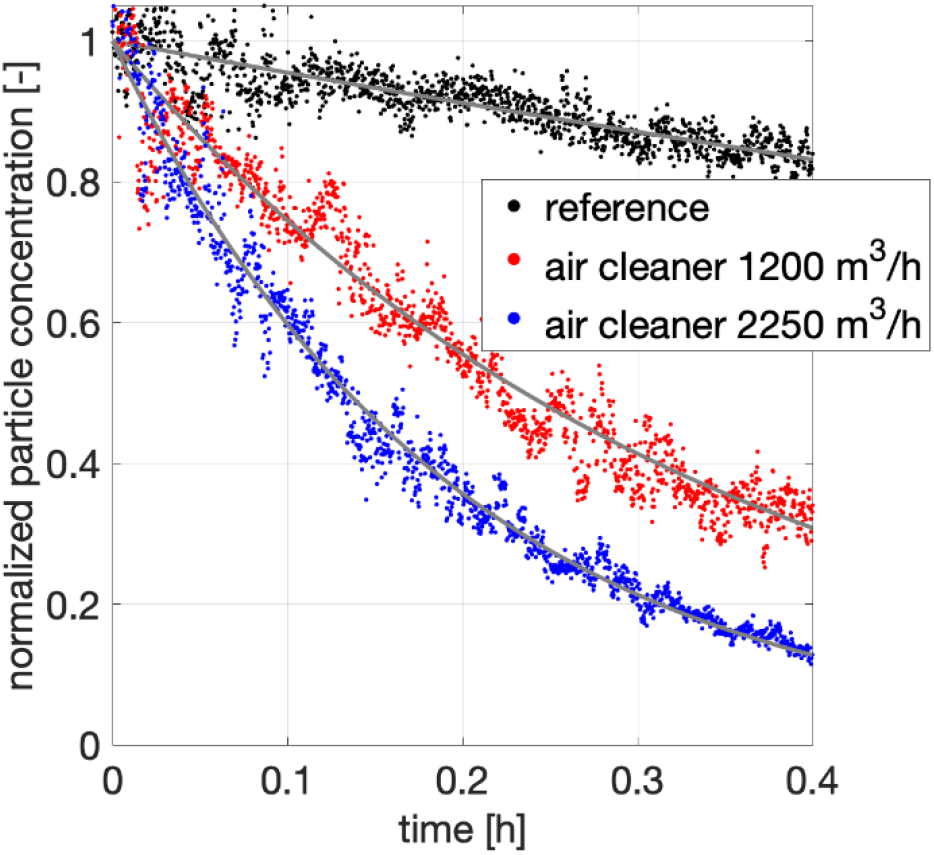
Normalized particle concentrations over time for the reference measurement and the room air cleaner at two different volume flows.

For a quantitative comparison, the decay rates determined for different configurations are shown in Table 1. “Reference” indicates the condition with closed windows and closed door as well as turned off HVAC system and without operation of the mobile room air cleaner. Due to window and door leakage, this value is not 0, but very low, as is completely normal for modern, well-insulated buildings. With the HVAC system turned on, the value of *k* is close to 6 (operating at 73% capacity (1200 m^3^/h) with 100% outside air), which is recommended by Harvard T.H. Chan School of Public Health (2021) and seems to be a reasonable value from a risk assessment, see Müller et al. (2020).

**Table 1:** Decay rates determined for different configurations.

| Configuration | Decay rate $k$ [1/h] |
| --- | --- |
| Reference | 0,5 |
| HVAC (73% or 1200 m<sup>3</sup>/h) | 5,7 |
| Window ventilation (only window 1 open) | 1,7 |
| Window ventilation (window 1 and 2 open) | 8,8 |
| Room air cleaner, 1200 m<sup>3</sup>/h | 2,9 |
| Room air cleaner, 2250 m<sup>3</sup>/h | 5,4 |

Window ventilation is shown in comparison. For this purpose, only one or both windows were fully opened. If the HVAC system is turned off and only window 1 is opened (shock ventilation), this results in a value of 1.7 for *k*. This value depends on the size of the window opening, the wind conditions in front of the window and the temperature difference between inside and outside (Etheridge and Sandberg (1996)). Since the latter varies greatly, this value is not constant. If both windows are opened (cross-ventilation), the value is 8.8. On the day of the experiment, the outdoor environment was windy with an average wind speed of around 10−15 km/h (clear movement of the trees can be seen). When there is no wind, the *k* value is significantly lower and primarily determined by the temperature difference between indoors and outdoors. With a mobile room air cleaner, the decay constant is much greater than with a permanently opened single window. However, the performance of the HVAC system outperforms the mobile air purifier for the same volume flow. This is basically due to the distributed openings for the in- and outflow, which makes the room air cleaning more efficient. To obtain a comparable decay rate, the volume flow of the room air cleaner had to be almost doubled. However, in this case inefficiencies become significant as the concept of mixing ventilation does not hold anymore perfectly.

### 4. Conclusion

The measurements carried out in room 2431 of building 033 show a very good efficiency of the HVAC system. In the tests, an air exchange rate of almost 6 was achieved. To ensure that this value is always achieved during operation, the system must be operated at least at the 73% performance level with 100% outside air and no control (e.g. based on the CO_2_ content) may be used that could lead to a reduction in the air supply. Some of the windows in building 033 have contact switches. When the windows are opened, the HVAC system in the respective room is automatically switched off in order to save energy. Thus, in this case, ventilation through the HVAC system and window ventilation do NOT add up, but only ventilation through windows takes place. As the window ventilation is less effective than the HVAC system, windows should stay closed during class within the pandemic.

In the studies conducted here, the cross-ventilation through both windows has a slightly higher air exchange rate than that of the HVAC due to the strong wind outside the building. Other outdoor environments may result in even higher air exchange rates, but in most cases they tend to be lower. In addition, at the air change rate determined in this experiment, both windows would have to actually be open about 2/3 of on hour to achieve similar efficiency to the HVAC per hour. The opening and closing of the windows would have to occur at regular short intervals to avoid an increase in the virus load in the enclosed space. Out of 15 minutes, they would need to be open for 10 minutes and closed for 5 minutes to achieve similar efficiency as the HVAC. In the cold months this leads to a strong temperature decrease in the room, and unpleasant conditions due to drafts, so that compared to the concept of window ventilation, the HVAC system is clearly to be preferred. Frequent opening of windows also disrupts lessons and requires additional work. Furthermore, it is only allowed to open windows when it is safe to do so.

Mobile air cleaners can achieve a comparable cleaning performance as the HVAC system, but the volume flow must be increased, since the HVAC system works more efficiently due to the distributed supply and exhaust air openings. Due to the increased volume flow, noise generation can be significant. To counter this problem, the use of two mobile air cleaners is recommended. In this way, filter performance comparable to that of the HVAC system can be achieved with relatively low noise emissions and a distributed air exchange is also possible, making the air purification more efficient. However, the reduction in noise is accompanied by an increase in costs. It must be emphasized, however, that in the rarest cases buildings are equipped with such efficient HVAC systems. The retrofitting of such HVAC system would mean a multiple of the purchase costs of mobile room air cleaners and a lengthy modernization of the building. Therefore, mobile air cleaners are a fast and cost-effective solution, which, in contrast to free ventilation, continuously provide a high level of air filtration, regardless of the size of the windows, wind and temperature conditions, and without interrupting the work by regularly opening and closing the windows and without unpleasant temperature and draft conditions in the room.

Based on this analysis, it can be concluded that a reduction of the virus load in the room can be realized very efficiently with a highly modern HVAC system. Therefore, these systems should be used when they are available. However, it is important to note that due to the hazardous nature of the virus, these systems should operate with 100% outside air and supply at least 6 times the room volume per hour to the room. It should be noted, however, that this air exchange rate can cause mucous membranes to dry out in the winter, making infections more likely. HVAC systems are also very quiet and relatively energy efficient if they use heat recovery.

Ventilation via windows can also lead to an efficient reduction of the virus load in the case of cross-ventilation. However, it should be noted that cross-ventilation must take place frequently and over long periods of time in order to achieve a comparable result to the HVAC system. Therefore, efficient cross-ventilation is always associated with a certain cooling of the room. This reduces the comfort and flu infections increase due to the decreasing humidity. Furthermore, the mucous membranes dry out. In the experiment, shock ventilation proved to be insufficient. Even with one-sided continuous ventilation, the air exchange rate achieved is not sufficient to significantly reduce the risk of infection. If significantly more windows could be opened, then the ventilation result would also improve, but then the temperature decrease in the room would be strong, which in turn would reduce the ventilation efficiency. However, more or larger windows which can be opened are often not present in modern buildings, because they are not required by law. Windows today serve to bring daylight into the room, but not to provide strong ventilation. That’s why most windows cannot be opened. For this reason, the one-sided window ventilation is only in rare cases practically suitable to reduce the virus load as the HVAC system is able to do. Furthermore, window ventilation is often associated with significant noise pollution, as outside noise from the street or the surrounding area enters the rooms. In addition, fine dust and pollen enter the room through open windows.

Room air cleaner work in recirculation mode and therefore no temperature effects occur. Only the CO_2_ increase and other vapors must be eliminated by window ventilation or special room air cleaners with exhaust air supply can be used (Kähler and Hain (2021)). However, these ventilation intervals are very large compared to the ventilation intervals required by window ventilation to effectively reduce the risk of infection. In principle, room air cleaners can achieve a comparable filtration performance as highly efficient air handling units, but not as quietly. When using several mobile air purifiers, the noise emission can be reduced, but at the expense of the purchase costs.

Finally, it should be noted that HVAC systems, room air cleaners and also ventilation via windows can only reduce the concentration of aerosol particles in the room. Thus, these measures are suitable for reducing the indirect infection risk due to a high viral load in the room. To reduce direct infections due to prolonged conversations over short distances, sufficiently large distances must be maintained, transparent protection walls must be installed, or mouth-nose covering must be worn. Hand hygiene should also be consistently observed in lecture halls to minimize the possibility of smear infections as well.

### Remark

The company Trotec GmbH (Heinsberg, Germany) financially supported the investigations. The room air cleaner of the type TAC V+ was provided by Trotec on loan free of charge for the investigations. This support has no effect on the investigations or results.

## Data Availability

Data can be provided upon request.

